# Clinical Utility Study of EsoGuard® on Samples Collected with EsoCheck® as a Triage Test for Endoscopy to Identify Barrett’s Esophagus – Interim Data of the First 275 Subjects

**DOI:** 10.1101/2023.08.31.23294916

**Authors:** Dan Lister, Andy Fine, Shail Maheshwari, Paul S. Bradley, Victoria T. Lee, Brian J. deGuzman, Suman Verma, Lishan Aklog

## Abstract

**Background:** Barrett’s Esophagus (BE) is the only known precursor for esophageal adenocarcinoma (EAC), a highly lethal malignancy which has had increasing incidence in Western populations over the last 40 years. Recommendations are for endoscopic screening of patients with multiple risk factors for BE, however most eligible patients are not undergoing such evaluation, or failing to be referred, leading to most patients with EAC being diagnosed without an existing BE diagnosis. EsoGuard® (EG) is a commercially available biomarker test for detection of BE, and when used to analyze cells collected non-endoscopically with EsoCheck® (EC), may serve as an easily accessible and well-tolerated diagnostic tool that has been recognized by the ACG and AGA as a reasonable alternative to screening endoscopy. The aim of this study was to evaluate the clinical utility of EG as a triage test for upper endoscopy in the diagnose BE in real world use.

**Methods:** We present the first data snapshot from a multi-center, observational trial evaluating the **CL**inical **U**tility of **E**soGuard (CLUE) among physicians who have adopted the technology into their clinical practice. At the time of data snapshot, four centers had contributed to enrollment of 275 subjects between February 23, 2023, to July 28, 2023. Participating centers followed their own standard practices for determining whom to test with EG on cells collected with EC and subsequent management of the patient following results. Demographics, risk factors, test results, and subsequent management decisions were collected and analyzed. The clinical utility of the technology was evaluated based on the impact of the EG test results on the ordering physician’s decision to refer or not refer a patient for further endoscopic evaluation.

**Results:** Among 275 subjects contributing data for analysis, the average age was 61.9 years, and there was a similar distribution among males and females. Eighty-nine-point seven percent (89.7%) reported a history of chronic GERD, and 73.8% had GERD plus an additional 3 BE risk factors (i.e., ACG screening cohort). 232 subjects had EG results documented at the time of data analysis, among which 229 also had a physician decision on endoscopy referral. Total EG positivity rate was 29.3% (68/232) and 65.5% (152/232) were negative; the positive agreement between positive EG results and referral for endoscopy was 100%. The negative agreement between a negative EG result and non-referral for endoscopy was 99.3%. The overall concordance between EG result and endoscopy referral was 98.8%. This did not substantially differ between the ACG screening cohort compared to others.

**Conclusions:** Data from the first snapshot of the CLUE study demonstrates physicians ordering EC/EG in the commercial setting are reliably utilizing EG results as a triage tool to guide referrals for endoscopic evaluation of BE. Physicians always refer EG(+) individual for additional endoscopic evaluation, whereas EG(-) subjects are consistently being spared an invasive test.

## Introduction

Barrett’s Esophagus (BE) is a metaplastic condition of the lower esophagus and is the only known precursor for esophageal adenocarcinoma (EAC), a malignancy which has had increasing incidence in Western populations over the last 40 years. [1] Experts and the general medical community identify the hallmark of BE as the presence of intestinal metaplasia i.e., the replacement of normal squamous epithelium with specialized columnar epithelium with intestinal-type goblet cells.[2] Screening for BE and surveillance for those diagnosed with disease is supported by multiple societal guidelines because contrary to the lethality of EAC, BE can be successfully treated using a number of endoscopic eradication therapies which achieve complete disease eradication in over 80-90% of treated patients. [3-5] Even with EAC, there is substantial improvement in survival if identified in the earliest stages, although this is infrequent as most patients present with dysphagia - by which time the cancer is usually advanced.[6] [7] As such, the underlying goal of BE screening strategies is to reduce EAC mortalities by diagnosis in the pre-neoplastic stage followed by either surveillance (non-dysplastic BE) or treatment (for dysplasia) to effectively halt disease progression.[4]

The diagnosis of BE is most frequently made when patients with refractory or severe GERD symptoms are found to have ≥1cm of “salmon colored mucosa” during upper endoscopy with presence of goblet cells on biopsy, but this approach to screening has several limitations. First, up to 44% of the population in some Western countries have GERD,[8] and when evaluating the incidence of other common risk factors (i.e., male sex, age >50 years, white race, etc.) it may not be realistic to perform screening endoscopy on everyone who meets criteria for elevated disease risk. Additionally, many patients with GERD utilize acid suppressive medications (recommended as part of disease management in published guidelines and from expert panels),[9] [10] and experience reasonable to good symptom control and may not seek or be referred for endoscopic evaluation, therefore BE in this population would continue to be missed. Unfortunately, while symptom control and reduced incidence of erosive esophagitis and peptic strictures are a benefit of acid suppressive medications such as proton pump inhibitors (PPIs), evidence suggests they do not reduce the risk of developing dysplasia or EAC in patients with BE.[11, 12] Clearly, better strategies for more widespread and earlier disease detection must be sought. One option is a two-step approach: first would be an easily accessible and non-invasive triage test to identify patients with high probability of having disease, followed by the more invasive confirmatory test which also allows disease staging (i.e., endoscopy with biopsies).

EsoGuard® (EG) is a commercially available biomarker assay that when performed on esophageal mucosal cells sampled using the non-endoscopic, balloon based EsoCheck® (EC) device, offers a minimally invasive alternative to upper endoscopy for initial detection of patients with BE. This is an accepted strategy recognized by both the American College of Gastroenterology (ACG) and American Gastroenterological Association (AGA).[4, 13] The goal of the ongoing, multicenter, prospective **CLUE** study is to capture real-world data from the commercial use of EG and evaluate the impact of test results on health care provider’s decision-making. CLUE focuses on patients with multiple risk factors for disease that meet either ACG or (at minimum) AGA recommendations for BE screening and are at elevated risk for disease compared to the general population. The analysis presented here is for the first 275 subjects enrolled into the study and for whom clinical utility data are available.

## Methods

To evaluate the utility of EG as a tool in the diagnosis of Barrett’s esophagus, patient demographics, risk factors, EG results, and provider management decisions were recorded and analyzed in a prospective, multi-center, observational study (***CL****inical* ***U****tility Study of* ***E****soGuard® on Samples Collected with EsoCheck® as a Triage Test for Endoscopy to Identify Barrett’s Esophagus* - **CLUE**). Because EG is intended as a triage test and upper endoscopy (with or without biopsies) deemed the ‘gold standard’ confirmatory test for diagnosis of BE, the key management decision captured in this study is the ordering physician’s decision whether to refer a patient for endoscopy based on his/her EG result.

Study sites are those in which the physicians have adopted the use of EC/EG technology into his/her standard practice, and therefore are not deviating from usual care as part of study conduct. The first site was initiated on 23-February-2023, and enrollment will continue until a target of 500 evaluable subjects has been reached. We present here the interim data collected through 28-July-2023 from the first 275 subjects enrolled across four study sites.

The study was conducted according to the guidelines of the Declaration of Helsinki and approved by the WCG Institutional Review Board (IRB tracking number 20222402). All participating individuals signed informed consent prior to EC and collection of any study information.

### EsoCheck® and EsoGuard® (EC/EG)

EsoCheck® is an FDA 510K cleared, non-endoscopic device designed for the circumferential, targeted collection and retrieval of surface cells from the esophagus. The unique, balloon-capsule technology allows for easy swallowing, non-traumatic cell sampling, and dilution protection of the specimen upon retrieval of the device through the upper esophagus and oropharynx (**Figure 1**). It is cleared for use in the general population of individuals 12 years or older. Cell collection is easy to perform in any office setting without sedation or specialized equipment, is well-tolerated, and usually takes less than 5 minutes.

**Figure 1.**
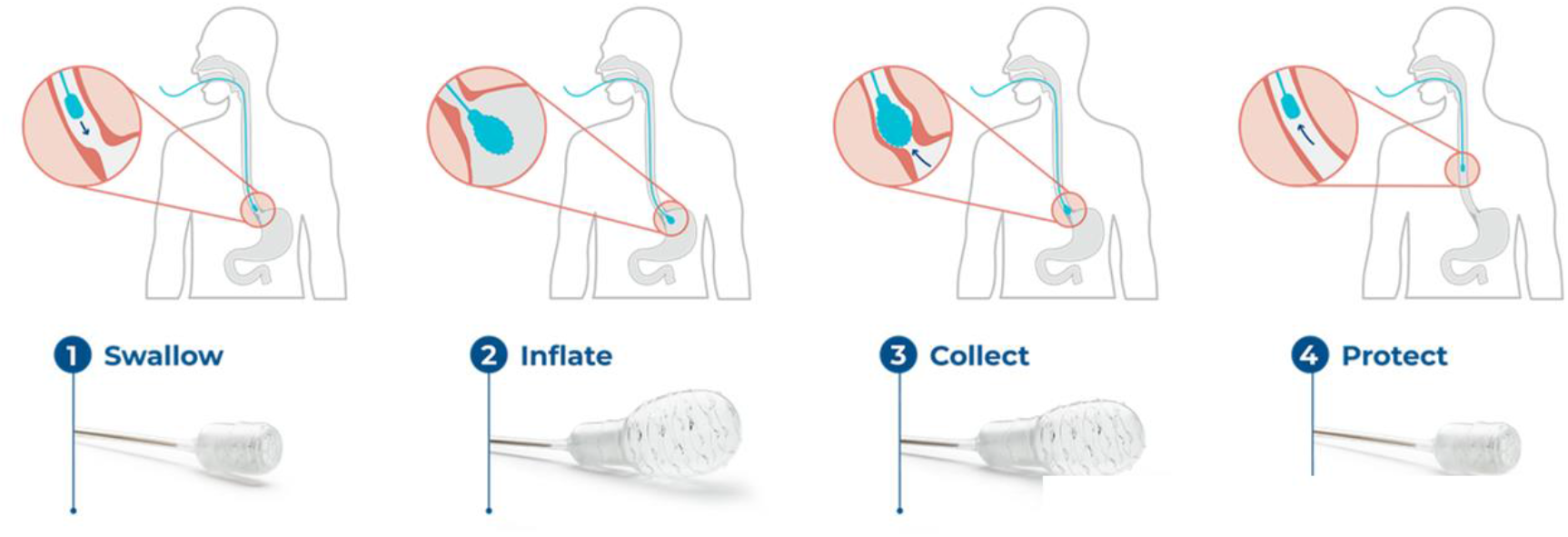
EsoCheck Cell Collection Process.

EsoGuard® (EG) is a laboratory developed test (LDT) performed in a Clinical Laboratory Improvement Amendment (CLIA) certified and College of American Pathologists (CAP) accredited Central Lab that utilizes a set of genetic assays and algorithms which examine the presence of cytosine methylation at 31 different genomic locations on the vimentin (VIM) and Cyclin-A1 (CCNA1) genes. EG has been clinically validated in a developmental study published in 2018 and shown to have a >90% sensitivity and >90% specificity in non-endoscopic detection of Barrett’s Esophagus (BE) or esophageal adenocarcinoma (EAC).[14] EG results are reported in a binary fashion (positive or negative) indicating presence or absence of sufficient methylation changes to suggest diagnosis of BE or disease along the BE progression spectrum. The patient and provider are notified of Quantity Not Sufficient (QNS) if the cell sample has insufficient DNA for EG analysis. Contaminated or otherwise unevaluable samples are reported as such, and the patient has the option to re-test.

### Study Population

Patients eligible for study participation are those whom a) their provider has made the independent clinical decision that a subject is at increased risk for disease compared to the general population and b) determined medical necessity to test a subject for BE using EC/EG (**Table 1**). “Increased risk” is defined within the study as patients with ≥3 established risk factors, as described by the ACG and AGA in their 2022 BE screening guidelines and 2022 clinical practice updates, respectively.[4, 13] These include male sex, white (Caucasian, non-Hispanic) race, chronic gastroesophageal reflux disease (GERD), history of tobacco smoking, obesity, age >50 years old, and/or family history of BE or EAC in a first degree relative. **Figure 2** provides a study schematic demonstrating the flow of the patient journey and data collected during the study.

**Table 1.** Inclusion and Exclusion Criteria.

| Inclusion Criteria | Exclusion Criteria |
| --- | --- |
| Signed informed consent | Inability to provide written informed consent or participate in the required follow up |
| Individuals in whom the decision has been made to test for BE using EC/EG | Individuals falling outside the indicated population defined by the EC device Instructions for Use (IFU) |
| Individuals who at minimum have three (3) or more established risk factors for BE or EAC:<br>Chronic GERD (defined as $\geq 5$ years of frequent symptoms)<br>Male sex<br>White race (Caucasian, non-Hispanic)<br>Age $>50$ years old<br>History of tobacco smoking<br>Obesity (defined as BMI $\geq 30$)<br>Family history of BE or EAC in a first degree relative | Individuals not meeting criteria for BE screening |

**Figure 2.**
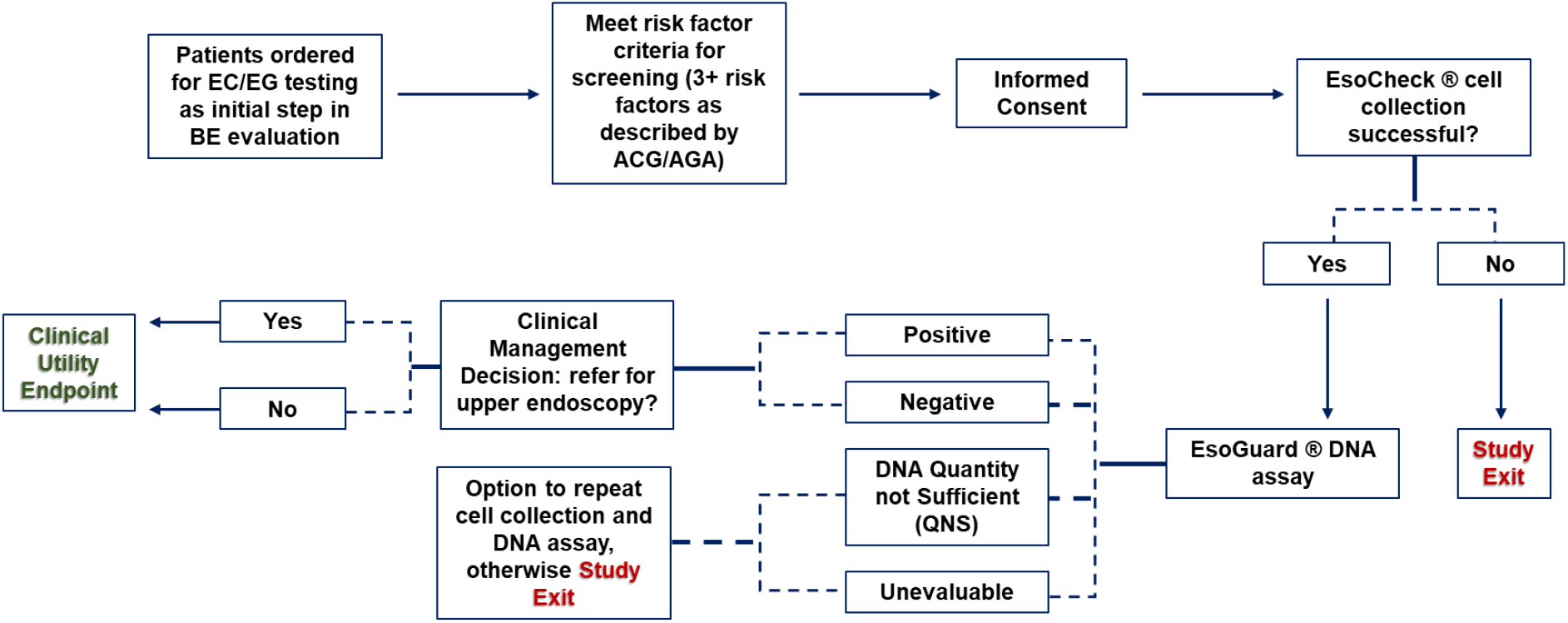
Study Schematic of CLUE. (**CL**inical **U**tility Study of **E**soGuard® on Samples Collected with EsoCheck® as a Triage Test for Endoscopy to Identify Barrett’s Esophagus)

### Statistical Analysis

Subjects unable to successfully swallow the EC device could not contribute cellular DNA for EG analysis; these subjects are included in the summary of enrollment demographics and risk factors, but do not contribute to the clinical utility endpoint. Similarly, subjects with QNS or cell samples deemed unevaluable on EG were included in overall data analysis but did not contribute to the primary clinical utility endpoints.

The primary clinical utility endpoints of this study included positive and negative agreement between EG results and endoscopy referral patterns, and the overall concordance between the EG result and provider decision for endoscopy referral. Positive agreement was calculated as the % of patients with EG positive (+) results who are referred for confirmatory upper endoscopy; negative agreement was calculated as the % of patients with EG negative (-) results who are not referred for any upper endoscopy.

Continuous variables were summarized using the number of observations (n), mean, standard deviation (SD), median, minimum, and maximum, along with total number of patients contributing values. Categorical variables were described by frequency of counts and percentages. The total number of applicable subjects (N) were used as the denominator for percent calculations unless stated otherwise within a table footnote. Binomial exact two-sided 95% confidence interval were calculated wherever relevant.

## Results

At the time of data snapshot, four clinical sites - each with a single, primary participating physician - had enrolled patients into CLUE. Two of the four participating physicians are primary care providers/internists, one a foregut surgeon, and the fourth a gastroenterologist. The participating foregut surgeon is also an endoscopist. A total of 279 subjects signed informed consent for study participation, however four (4) were noted after consent to not appropriately meet inclusion criteria and were withdrawn early from participation resulting in 275 subjects contributing data for analysis.

### Subject Characteristics and Risk Factors

Subject baseline characteristics and BE risk factors are summarized in **Table 2**. Four of the enrolled subjects were pending entry of demographic information at the time of data snapshot The mean age was 61.9 years (SD 12.6 years), with a relatively equitable distribution among male vs. female sex. Most subjects (76.0%) were of White race (i.e., Caucasian non-Hispanic), and nearly 90% had a history of chronic gastroesophageal reflux disease (GERD). Among the cohort with GERD, the average duration of the condition was over 14 years, speaking to the chronicity, and most used acid suppressive medications with good symptom response. Most other BE risk factors were also well-represented, although a positive family history of BE/EAC in a first degree relative was infrequent (<3%).

**Table 2.**
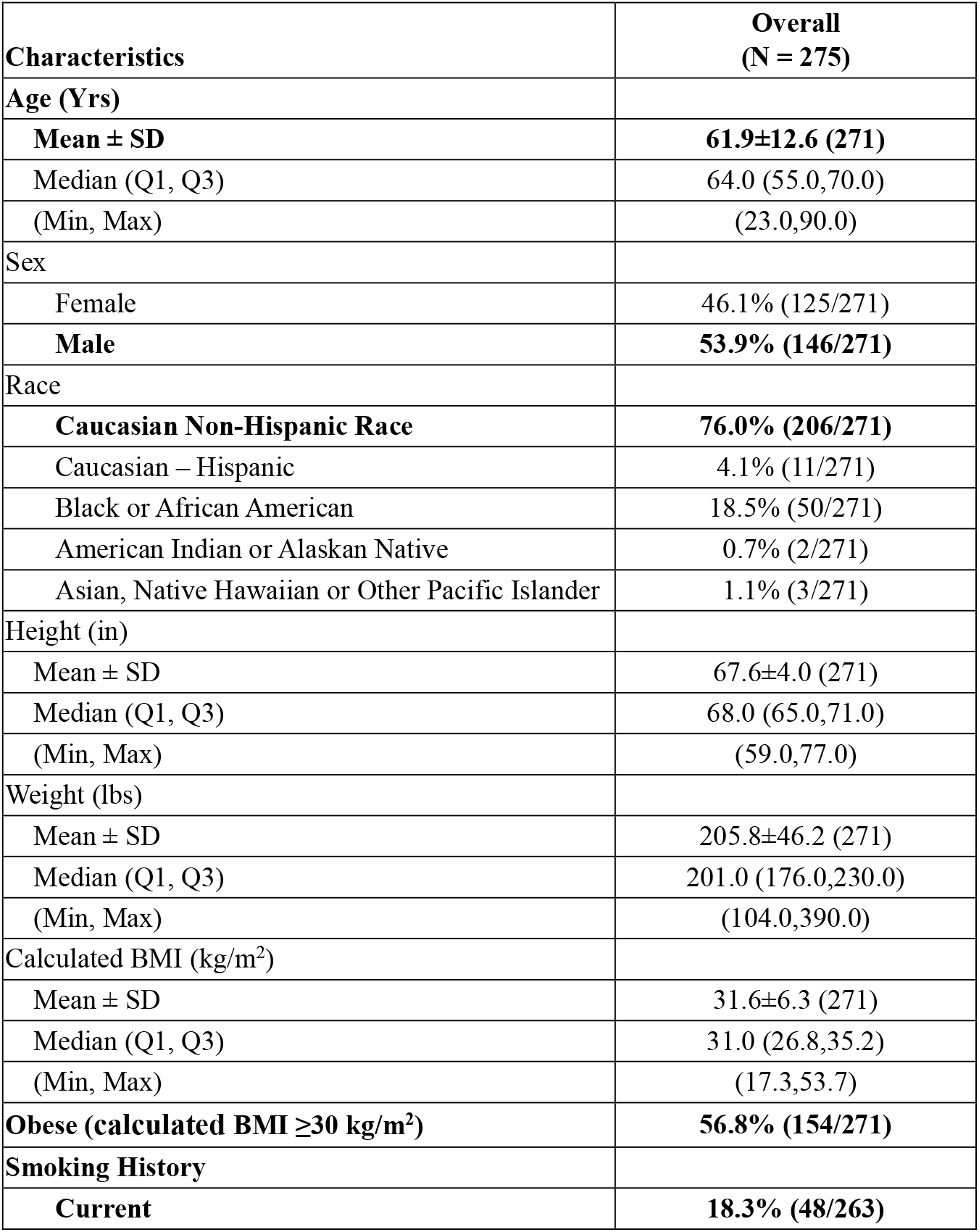
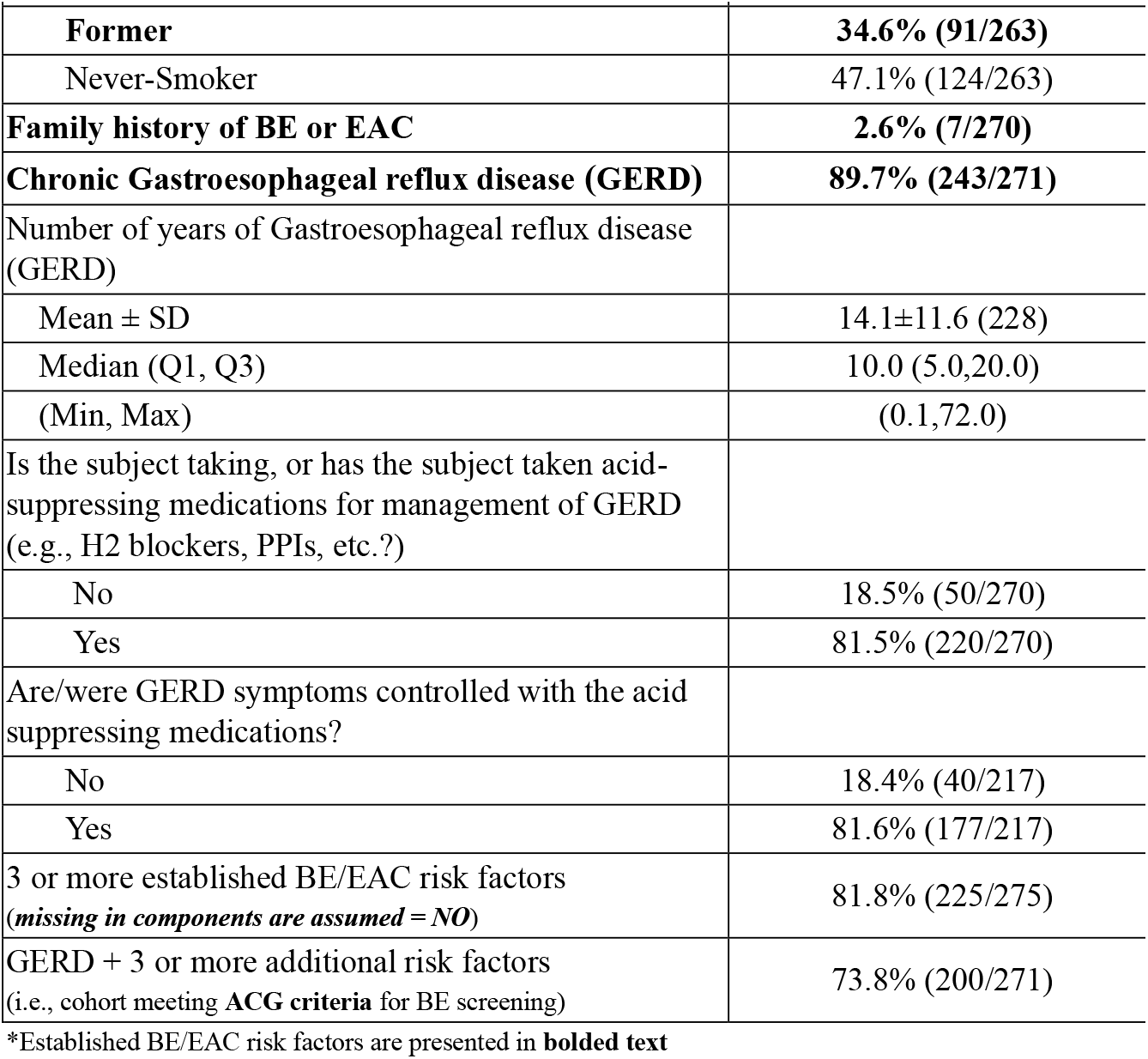
Subject Baseline Characteristics and BE/EAC Risk Factors.

| **Characteristics** | **Overall**<br>**(N = 275)** |
| --- | --- |
| **Age (Yrs)** |  |
| **Mean ± SD** | **61.9±12.6 (271)** |
| Median (Q1, Q3) | 64.0 (55.0,70.0) |
| (Min, Max) | (23.0,90.0) |
| **Sex** |  |
| Female | 46.1% (125/271) |
| **Male** | **53.9% (146/271)** |
| **Race** |  |
| **Caucasian Non-Hispanic Race** | **76.0% (206/271)** |
| Caucasian – Hispanic | 4.1% (11/271) |
| Black or African American | 18.5% (50/271) |
| American Indian or Alaskan Native | 0.7% (2/271) |
| Asian, Native Hawaiian or Other Pacific Islander | 1.1% (3/271) |
| **Height (in)** |  |
| Mean ± SD | 67.6±4.0 (271) |
| Median (Q1, Q3) | 68.0 (65.0,71.0) |
| (Min, Max) | (59.0,77.0) |
| **Weight (lbs)** |  |
| Mean ± SD | 205.8±46.2 (271) |
| Median (Q1, Q3) | 201.0 (176.0,230.0) |
| (Min, Max) | (104.0,390.0) |
| **Calculated BMI (kg/m<sup>2</sup>)** |  |
| Mean ± SD | 31.6±6.3 (271) |
| Median (Q1, Q3) | 31.0 (26.8,35.2) |
| (Min, Max) | (17.3,53.7) |
| **Obese (calculated BMI ≥30 kg/m<sup>2</sup>)** | **56.8% (154/271)** |
| **Smoking History** |  |
| **Current** | **18.3% (48/263)** |

**Table 2. Subject Baseline Characteristics and BE/EAC Risk Factors**
| Characteristics | Overall<br>(N = 275) |
| --- | --- |
| **Former** | **34.6% (91/263)** |
| Never-Smoker | 47.1% (124/263) |
| **Family history of BE or EAC** | **2.6% (7/270)** |
| **Chronic Gastroesophageal reflux disease (GERD)** | **89.7% (243/271)** |
| Number of years of Gastroesophageal reflux disease (GERD) |  |
| Mean ± SD | 14.1±11.6 (228) |
| Median (Q1, Q3) | 10.0 (5.0,20.0) |
| (Min, Max) | (0.1,72.0) |
| Is the subject taking, or has the subject taken acid-suppressing medications for management of GERD (e.g., H2 blockers, PPIs, etc.?) |  |
| No | 18.5% (50/270) |
| Yes | 81.5% (220/270) |
| Are/were GERD symptoms controlled with the acid suppressing medications? |  |
| No | 18.4% (40/217) |
| Yes | 81.6% (177/217) |
| 3 or more established BE/EAC risk factors<br>(missing in components are assumed = NO) | 81.8% (225/275) |
| GERD + 3 or more additional risk factors<br>(i.e., cohort meeting **ACG criteria** for BE screening) | 73.8% (200/271) |

\*Established BE/EAC risk factors are presented in **bolded text**

Most of the study population met ACG guideline criteria for endoscopic BE screening, at 73.8%. A small number of individuals had incomplete data on their risk factors.

### EsoCheck Cell Collection

EC cell collection was performed in accordance with the device’s instructions for use (IFU, available upon request from https://www.luciddx.com/esocheck) Of all enrolled subjects, EsoCheck cell collection information was available for 272, among which 96.3% successfully completed cell collection (**Table 3**). The subjects unable to tolerate the cell collection (3.7%, 10/272) were exited from the study early.

**Table 3.**
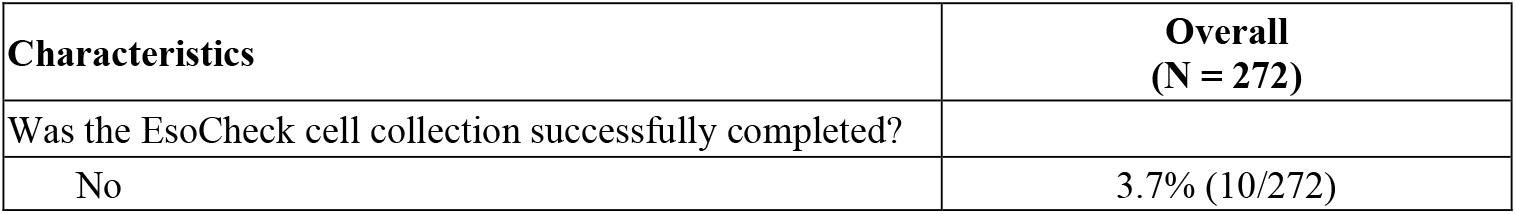
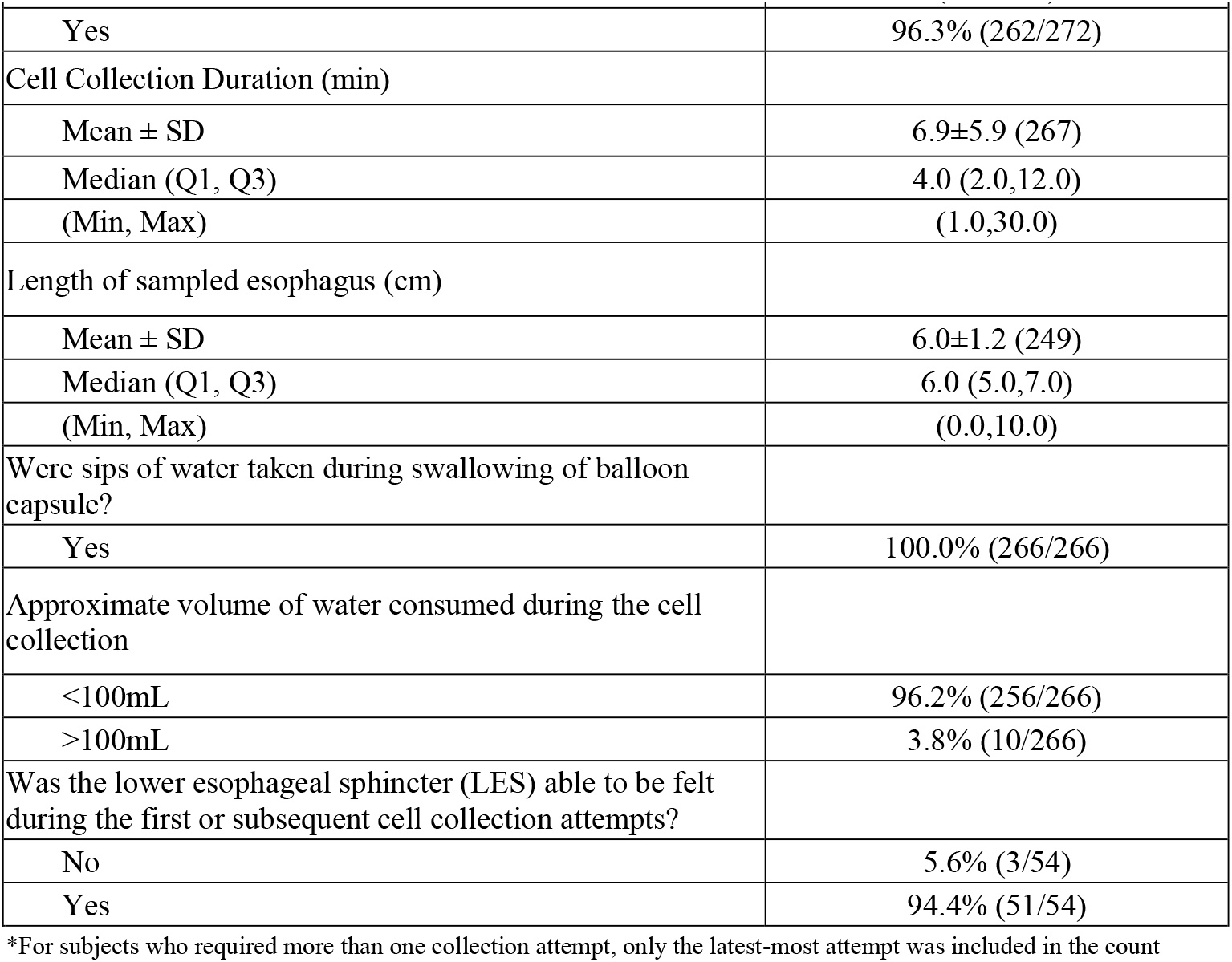
EsoCheck Cell Collection Characteristics.

Median duration of the cell collection process was 4min (119/267 subjects (44.5%) completed the cell collection in 3min or less); the fastest cell collections occurred in one minute, and a maximum collection time of 30min was seen in one individual who required several attempts to swallow the EsoCheck device. All subjects utilized small sips of water to facilitate device swallowing, with most requiring <100mL. Mean length of sampled esophagus was 6cm, which is consistent with the EsoCheck Instructions For Use (IFU).

### EsoGuard Results and Clinical Utility Evaluation

Of the 272 subjects with EsoCheck cell collection information, 242 had received EG results by the time of data snapshot, although only 232 were documented in the study database. Among those 229 also had a documented management decision from their ordering physician regarding referral for upper endoscopy (**Table 4A**). Just under 30% of the EG results returned positive (29.3%, 68/232) and 65.5% (152/232) returned negative. Eight subjects (3.4%) had insufficient DNA quantity in their cell samples for EG analysis (QNS), and four (1.7%) cell samples were unevaluable due to other factors (e.g., contamination).

**Table 4A.** Summary of EsoGuard Results and Physician Decisions on Endoscopy Referral.

| Characteristics | Overall<br>(N = 272*) |
| --- | --- |
| Was the EsoGuard assay completed on the collected cell sample? |  |
| No** | 4.1% (10/242) |
| Yes | 95.9% (232/242) |
| EsoGuard assay result: |  |
| NEGATIVE | 65.5% (152/232) |
| POSITIVE | 29.3% (68/232) |

| Characteristics | Overall<br>(N = 272*) |
| --- | --- |
| QUANTITY NOT SUFFICIENT (QNS) | 3.4% (8/232) |
| UNEVALUABLE | 1.7% (4/232) |
| Was the subject referred for upper endoscopy? |  |
| No | 69.4% (159/229) |
| Yes | 30.6% (70/229) |
| Provide the reason for referring or not referring the patient for an endoscopy: |  |
| Due to NEGATIVE EsoGuard Result | 65.9% (151/229) |
| Due to POSITIVE EsoGuard Result | 29.7% (68/229) |
| OTHER | 4.4% (10/229) |
| Other, please specify: |  |
| Endoscopy required for evaluation of reflux surgery§ | 10.0% (1/10) |
| Patient refused endoscopy↓ | 10.0% (1/10) |
| QNS or unevaluable EsoGuard result – pending repeat test; no endoscopy referral until further results available | 50.0% (5/10) |
| Unevaluable EsoGuard result – subject referred for endoscopy rather than repeat test, given his/her risk factors | 10.0% (1/10) |
| QNS EsoGuard result – subject not warranted for endoscopy without a positive result | 20.0% (2/10) |

\*All subjects who completed EsoCheck cell collection are included in this count, even if EsoGuard results have not yet been processed; average time from cell collection to results is 7-14 days; some results may also have been received by the ordering provider but not yet entered in the study database
\*\* Cell samples shipped to the Central Lab for analysis but for which EsoGuard results are still pending were reported here as “not completed” by some sites
§Subject had a negative EG result and was scheduled for upper endoscopy for non-screening purposes
↓Subject had a positive EG result and was referred for endoscopy, but refused scheduling of the procedure

Just over 30% of subjects (70/229) were referred for upper endoscopy following their EG results; the remainder were not. According to the investigators, the reason for over 95% of their endoscopy referral decisions was the positive (29.7%) or negative (65.9%) EG result (**Table 4A**).

EG results and their relationship to endoscopy referral decisions were evaluated by subject risk cohort (those either meeting ACG screening criteria or not) and presented in **Table 4B**. Three subjects with non-binary EG results (two QNS and one unevaluable) were pending endoscopy referral decisions. Two EG(+) subjects and one EG(-) subject with endoscopy referral decisions were missing complete risk factor and/or demographic information and therefore could not be classified into either the ACG vs. non-ACG cohorts; these subjects were excluded from counts within those cohorts but still contributed to analysis of the full study cohort. All subjects with EG(+) results were referred for confirmatory upper endoscopy. This was consistent across both risk cohorts. Only one subject with a negative EG result was referred for endoscopy, and one subject with an unevaluable result was also referred directly to endoscopy rather than repeating EC/EG.

**Table 4B.**
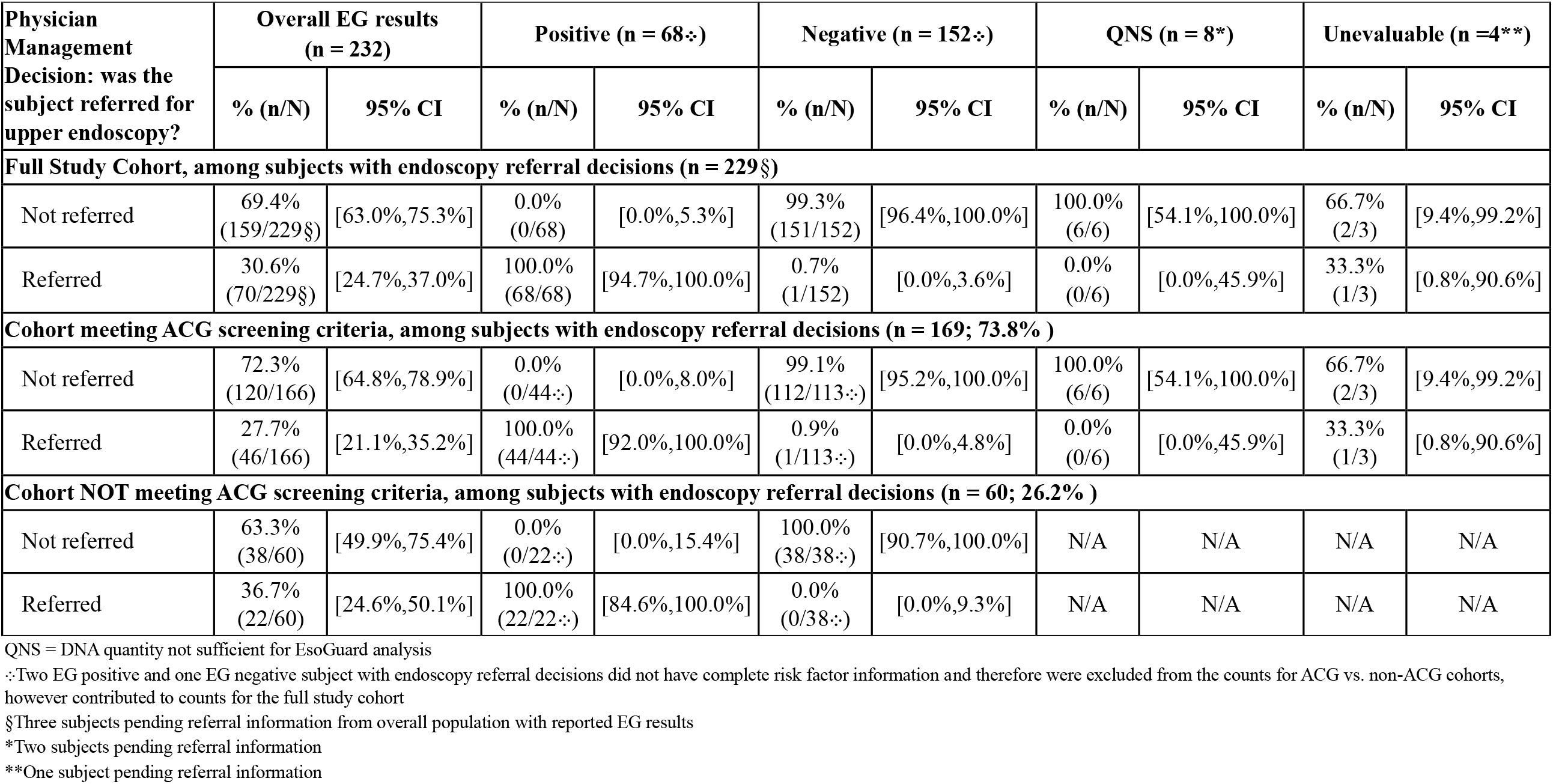
EsoGuard Results and Endoscopy Referral Decisions By Risk Cohort.

| Physician Management Decision: was the subject referred for upper endoscopy? | Overall EG results (n = 232) |  | Positive (n = 68§) |  | Negative (n = 152§) |  | QNS (n = 8*) |  | Unevaluable (n =4**) |  |
| --- | --- | --- | --- | --- | --- | --- | --- | --- | --- | --- |
|  | % (n/N) | 95% CI | % (n/N) | 95% CI | % (n/N) | 95% CI | % (n/N) | 95% CI | % (n/N) | 95% CI |
| **Full Study Cohort, among subjects with endoscopy referral decisions (n = 229§)** |  |  |  |  |  |  |  |  |  |  |
| Not referred | 69.4%<br>(159/229§) | [63.0%,75.3%] | 0.0%<br>(0/68) | [0.0%,5.3%] | 99.3%<br>(151/152) | [96.4%,100.0%] | 100.0%<br>(6/6) | [54.1%,100.0%] | 66.7%<br>(2/3) | [9.4%,99.2%] |
| Referred | 30.6%<br>(70/229§) | [24.7%,37.0%] | 100.0%<br>(68/68) | [94.7%,100.0%] | 0.7%<br>(1/152) | [0.0%,3.6%] | 0.0%<br>(0/6) | [0.0%,45.9%] | 33.3%<br>(1/3) | [0.8%,90.6%] |
| **Cohort meeting ACG screening criteria, among subjects with endoscopy referral decisions (n = 169; 73.8% )** |  |  |  |  |  |  |  |  |  |  |
| Not referred | 72.3%<br>(120/166) | [64.8%,78.9%] | 0.0%<br>(0/44:) | [0.0%,8.0%] | 99.1%<br>(112/113:) | [95.2%,100.0%] | 100.0%<br>(6/6) | [54.1%,100.0%] | 66.7%<br>(2/3) | [9.4%,99.2%] |
| Referred | 27.7%<br>(46/166) | [21.1%,35.2%] | 100.0%<br>(44/44:) | [92.0%,100.0%] | 0.9%<br>(1/113:) | [0.0%,4.8%] | 0.0%<br>(0/6) | [0.0%,45.9%] | 33.3%<br>(1/3) | [0.8%,90.6%] |
| **Cohort NOT meeting ACG screening criteria, among subjects with endoscopy referral decisions (n = 60; 26.2% )** |  |  |  |  |  |  |  |  |  |  |
| Not referred | 63.3%<br>(38/60) | [49.9%,75.4%] | 0.0%<br>(0/22:) | [0.0%,15.4%] | 100.0%<br>(38/38:) | [90.7%,100.0%] | N/A | N/A | N/A | N/A |
| Referred | 36.7%<br>(22/60) | [24.6%,50.1%] | 100.0%<br>(22/22:) | [84.6%,100.0%] | 0.0%<br>(0/38:) | [0.0%,9.3%] | N/A | N/A | N/A | N/A |

QNS = DNA quantity not sufficient for EsoGuard analysis
∴Two EG positive and one EG negative subject with endoscopy referral decisions did not have complete risk factor information and therefore were excluded from the counts for ACG vs. non-ACG cohorts, however contributed to counts for the full study cohort
§Three subjects pending referral information from overall population with reported EG results
\*Two subjects pending referral information
\*\*One subject pending referral information

Two hundred twenty subjects had a binary EG result (positive or negative) *and* a documented physician decision on endoscopy referral, thus contributing to analysis of the primary clinical utility endpoint (**Table 5**). The ***positive agreement*** was 100% in both the full study population, and the ACG screening cohort. ***Negative agreement*** was 99.3% in the full study population, and 99.1% in the ACG screening cohort. The overall concordance between EG result and endoscopy referral pattern was over 98% for both the full study population and the ACG screening cohort. When evaluating the clinical utility endpoint across participating study sites, all except one had 100% concordance.

**Table 5.** Primary Clinical Utility Endpoint(s) by Study Site.

| Analysis Set | Subjects with Binary EG Result | Positive Agreement (95% CI) | Negative Agreement (95% CI) | Concordance (95% CI) |
| --- | --- | --- | --- | --- |
| **Overall** | **220** | **100.0% (94.7%,100.0%)** | **99.3% (96.4%,100.0%)** | **98.9% (96.9%,100.0%)** |
| Site ID = 01 | 22 | 100.0% (29.2%,100.0%) | 94.7% (74.0%,99.9%) | 83.1% (51.1%,100.0%) |
| Site ID = 02 | 42 | 100.0% (75.3%,100.0%) | 100.0% (88.1%,100.0%) | 100.0% (100.0%,100.0%) |
| Site ID = 03 | 71 | 100.0% (81.5%,100.0%) | 100.0% (93.3%,100.0%) | 100.0% (100.0%,100.0%) |
| Site ID = 05 | 85 | 100.0% (89.7%,100.0%) | 100.0% (93.0%,100.0%) | 100.0% (100.0%,100.0%) |
| **ACG Cohort** | **157** | **100.0% (92.0%,100.0%)** | **99.1% (95.2%,100.0%)** | **98.4% (95.4%,100.0%)** |
| Site ID = 01 | 22 | 100.0% (29.2%,100.0%) | 94.7% (74.0%,99.9%) | 83.1% (51.1%,100.0%) |
| Site ID = 02 | 32 | 100.0% (66.4%,100.0%) | 100.0% (85.2%,100.0%) | 100.0% (100.0%,100.0%) |
| Site ID = 03 | 50 | 100.0% (71.5%,100.0%) | 100.0% (91.0%,100.0%) | 100.0% (100.0%,100.0%) |
| Site ID = 05 | 85 | 100.0% (89.7%,100.0%) | 100.0% (93.0%,100.0%) | 100.0% (100.0%,100.0%) |

## Discussion

Despite well-established criteria defining patients at increased risk for Barrett’s Esophagus and multiple published societal guidelines for screening, a significant diagnostic gap remains, as most patients who could benefit from testing are not being tested.[15] When different modalities for BE screening were reviewed and compared – including traditional endoscopy, transnasal endoscopy, video capsule endoscopy, and minimally invasive sampling devices (combined with analysis of cellular markers) – it was apparent that those with the highest diagnostic accuracy (i.e., endoscopy) were also associated with the lowest transportability and patient convenience or acceptance.[16] This supports the concept of a two-step process for improved BE diagnosis: the first step being a well-tolerated, highly sensitive, non-invasive triage test which enables greater accessibility for the large, at-risk population; the second step would be a confirmatory test (for triage ‘positive’ patients only) with high diagnostic accuracy but lower convenience – namely endoscopy with or without biopsy.

Patient triage via non-endoscopic testing strategies has accumulated increasing interest, with the most widespread literature available for CytoSponge, a swallowable sponge on a string, paired with immunohistochemistry (trefoil factor /TTF3).[17] Comparative modeling analyses have even shown that use of this diagnostic approach in primary care settings can be cost effective.[18] In China, balloon-based esophageal cell collection has been successful in supporting cytology screening for esophageal cancer.[19] Aside from being the only commercially available non-endoscopic esophageal cell collection device on the U.S market, advantages of EsoCheck (EC) compared to the other devices include the unique, balloon-capsule design, which allows targeted cell collection and specimen protection. Specifically for diagnosis of BE, a disease in which cellular changes originate and are localized to the distal esophagus, balloon inversion within the EC capsule after targeted collection in the distal esophagus, avoids cellular dilution and contamination as the device is removed through the upper esophagus and oropharynx. Additionally, as seen in CLUE, the EC cell collection process is fast, with a median cell collection time of only four minutes (note - the mean duration was skewed by the presence of one extreme outlier), and very well tolerated with less than four percent of patients unable to swallow the device; no patients reported complaints or complications to their physicians following the visit. This contrasts with sponge-based cell collection devices that take a minimum of 7 minutes for the gel capsule to dissolve in the stomach, and run the risk of string detachment.[17, 20] The EsoGuard (EG) biomarker assay, which utilizes next generation sequencing and validated algorithms to detect abnormal DNA methylation patterns, in turn has significant advantages over cytology. Unlike cytology, which requires highly trained experts to accurately classify cells, the EG assay is automated, easily scalable, and not subject to inter-observer variability. As seen in the CLUE data snapshot, binary EG test results were available in approximately 95% of patients, which remains well within standards set within the lab’s CLIA requirements.

Patients included within this CLUE interim analysis accurately represent the target BE testing population as described by GI society guidelines, namely patients with multiple risk factors - the majority of which have chronic GERD of long-standing duration. Over 80% of the chronic GERD patients in the study were on acid suppressive medications, 81.6% of which reported good symptom control and would therefore have been less likely to seek out or be referred for upper endoscopy for screening for BE. The observed EG positivity rate of 29.3% may seem high compared to reported BE prevalence rates of 5-15% cited in the literature, however this number is reasonable in the context of the higher risk study population (majority of subjects with 4 or more established BE risk factors). Published BE prevalence rates are also likely an under-reporting of true disease prevalence due to historically low rates of screening, leading to under-diagnosis.[21, 22] Indeed, literature shows that less than 20% of patients in the U.S who are diagnosed with EAC have any preceding diagnosis of BE, and only 10% of high risk individuals are undergoing endoscopic BE screening.[23, 24] This is likely multifactorial in nature, including the absence of specific symptoms associated with BE, poor patient understanding of their own disease risk, and fears around the discomfort or inconvenience of upper endoscopy.[25] Office-based, non-endoscopic testing with EC/EG can address these patient concerns by improving accessibility and minimizing invasiveness. Additionally, given the intended utility of EsoGuard as a high-sensitivity triage test, it is expected that the test positivity rate should be higher than true disease prevalence, so as not to risk missing any patients with disease.

Although the physician sample size was small, three different specialty types were represented, with no substantial differences in how EG results influenced their subsequent clinical decision-making. The positive agreement endpoint of 100%, negative agreement endpoint of 99.3%, and overall concordance (between EG result and endoscopy referral patterns) of 98.8% demonstrates that CLUE physicians were indeed utilizing EG as a triage test, with assay results consistently determining the next step in patient management. This is consistent with the physician’s own self-reporting, with >95% of their referral “reasons” being either a negative or positive EG result (**Table 4A**). This remained true even for the cohort of patients that met ACG guideline criteria for endoscopic screening. The ACG screening guidelines for BE are arguably some of the more stringent compared to those of other GI societies, given their requirement that all patients have chronic GERD (defined as five or more years of frequent symptoms) ***and*** three additional risk factors.[26] These patients could be clinically justified in proceeding straight to endoscopy for BE screening, however in all except one individual with negative EG results (112/113, 99.1%), triage with EG was able to save them from the more burdensome, uncomfortable, and higher-risk diagnostic procedure. The singular subject with a negative EG result who was sent for upper endoscopy did so for non-screening purposes; instead, the endoscopy was performed for pre-operative workup of planned anti-reflux surgery (**Table 4A** and **4B**). This clearly demonstrates provider confidence in negative EG results and its ability to rule out likelihood of BE in their patient(s).

The findings of clinical utility analysis are unsurprising, given that groups including the AGA and ACG have recognized non-endoscopic cell collection paired with assessment of DNA biomarkers as an acceptable approach to initial BE screening.[4, 13] While the focus of this manuscript was on how physicians utilize EG in their real-world patient management, we recognize that absence of patient outcomes may be deemed a limitation. However, the intent of technologies like EC/EG are to facilitate early diagnosis and increase patient and provider awareness of BE. The intent is not to change standards of care following diagnosis. There are clear guidelines on the appropriate management of patients with established disease, including surveillance and timing/indications for ablative therapy. These guidelines have been established with the intent of directly improving immediate and long-term patient outcomes.[4, 27, 28] However, after disease diagnosis, it is not within the scope of a triage test like EG or studies like CLUE to ensure patient or provider compliance with those guidelines. Some may argue that another limitation of this study is inference of any impact that EG results had on physician management decisions, rather than directly collecting information on change(s) to decision making. This was addressed in a recent protocol amendment and will be included in future analyses. Finally, the small number of enrolling sites (n=4) and participating physicians may also be deemed a study limitation; the number of sites and physicians is planned to double over the remainder of the study. Of note, despite the small number of investigators, as described previously, multi-disciplinary representation was still achieved.

In short, the early experience in CLUE appears to support EG as an effective triage test that can be used in both primary care and specialty settings to assist in the decision-making process for patients deemed at increased risk of BE. This approach could enable broad outreach and more consistent testing of increased-risk patients, while also focusing upper endoscopy resources on those patients with the highest pre-procedure probability of disease.

## Conclusions

Review of data from the first snapshot of the CLUE study demonstrates that physicians who have adopted EC/EG into their clinical practices are reliably utilizing EG as a triage tool for endoscopic evaluation of BE. Physicians always refer EG(+) individual for additional endoscopic evaluation, whereas EG(-) subjects are consistently being spared the more invasive test.

## Data Availability

All data produced in the present work are contained in the manuscript

